# Emulation of a target trial from observational data to compare effectiveness of Casirivimab/Imdevimab and Bamlanivimab/Etesevimab for early treatment of non-hospitalized patients with COVID-19

**DOI:** 10.1101/2022.02.04.22270143

**Authors:** V Mazzotta, A Cozzi Lepri, F Colavita, S Lanini, R Rosati, E Lalle, I Mastrorosa, C Cimaglia, A Vergori, Nazario Bevilacqua, Lapa Daniele, Mariano Andrea, Aurora Bettini, C Agrati, P Piselli, E Girardi, C Castilletti, AR Garbuglia, F Vaia, E Nicastri, A Antinori

## Abstract

**Objectives:** Comparative analysis between different monoclonal antibodies (mAbs) against SARS-CoV-2 are lacking. We present an emulation trial from observational data to compare effectiveness of Bamlanivimab/Etesevimab (BAM/ETE) and Casirivimab/Imdevimab (CAS/IMD) in outpatients with early mild-to-moderate COVID-19 in a real-world scenario of variants of concern (VoCs) from Alpha to Delta.

**Methods:** Allocation to treatment was subject to mAbs availability, and the measured factors were not used to determine which combination to use. Patients were followed through day 30. Viral load was measured by cycle threshold (CT) on D1 (baseline) and D7.

Primary outcome was time to COVID-19-related hospitalization or death from any cause over days 0-30. Weighted pooled logistic regression and marginal structural Cox model by inverse probability weights were used to compare BAM/ETE vs. CAS/IMD. ANCOVA was used to compare mean D7 CT values by intervention. Models were adjusted for calendar month, MASS score and VoCs. We evaluated effect measure modification by VoCs, vaccination, D1 CT levels and enrolment period.

**Results:** COVID19-related hospitalization or death from any cause occurred in 15 of 237 patients in the BAM/ETE group (6.3%) and in 4 of 196 patients in the CAS/IMD group (2.0%) (relative risk reduction [1 minus the relative risk] 72%; p=0.024). Subset analysis carried no evidence that the effect of the intervention was different across stratification factors. There was no evidence in viral load reduction from baseline through day 7 across the two groups (+0.17, 95% −1.41;+1.74, p=0.83). Among patients who experienced primary outcome, none showed a negative RT-PCR test in nasopharingeal swab (p=0.009) and 82.4% showed still high viral load (p<0.001) on D7.

**Conclusions:** In a pre-Omicron epidemiologic scenario, CAS/IMD reduced risk of clinical progression of COVID-19 compared to BAM/ETE. This effect was not associated with a concomitant difference in virological response.

## 1. Introduction

The widespread of severe acute respiratory syndrome coronavirus 2 (SARS-CoV-2) infection, causing coronavirus disease-2019 (COVID-19), continues to be a challenge for global public health. Monoclonal antibodies (mAbs) against SARS-CoV-2 have emerged as the strategy of choice for the treatment of early mild-to-moderate COVID-19 in outpatients at increased risk of clinical progression^1^ and, based on data from randomized clinical trials (RCT), Bamlanivimab/Etesevimab^2–3^, Casirivimab/Imdevimab^4–5^ and Sotrovimab^6^ received emergency use authorizations by the Italian Medicines Agency (AIFA)^7^.

In addition to RCT, data from observational cohorts^8–9–10–11^ have also confirmed the effectiveness of mAbs, but comparative analysis between different options available are lacking^12–13^. Considering the rapid epidemiological evolution, with the emergence of new Variants of Concern (VoCs) ^14–15–16^ that have been shown to escape^17^ the action of mAbs *in vitro*^18–19^ and *in vivo*^20^, real-life data about clinical impact and mAb comparison are useful to better clarify the scenario of currently existing drugs.

The aim of this analysis was to compare the clinical effectiveness of two mAb combinations, Bamlanivimab/Etesevimab (BAM/ETE) and Casirivimab/Imdevimab (CAS/IMD), in a real-life setting during a period in which the prevalent lineages in Italy were B.1.1.7 (Alpha) and B.1.617.2 (Delta), and B.1.529 (Omicron) was not yet circulating.

## 2. Methods

### 2.1 Monoclonal antibody access program and eligible patients

On March 2021, mAb administration program started at the National Institute for Infectious Diseases Lazzaro Spallanzani IRCCS in Rome, ruled by AIFA. Eligibility criteria included outpatients with a confirmed diagnosis of SARS-CoV-2 infection by an antigenic or molecular nasopharyngeal swab (NPS), mild or moderate symptoms of COVID-19 for 10 days or less and at least one of following risk factors for progression to severe disease: body mass index (BMI) ≥35, chronical peritoneal dialysis or hemodialysis, uncontrolled or complicated diabetes mellitus, primary or secondary immunodeficiency. Subjects who were 55 years and older were also eligible if they had any cardio-cerebrovascular diseases, Chronic Obstructive Pulmonary Disease (COPD) or other chronic respiratory diseases. Patients requiring hospitalization for COVID-19 or supplemental oxygen therapy were excluded. In June 2021^21^, AIFA expanded use of mAbs including all patients with one of the following: 65 years and older, BMI >30, any chronic renal impairment (including subjects undergoing peritoneal dialysis or hemodialysis), uncontrolled or complicated diabetes mellitus, any immunocompromising condition, cardio-cerebrovascular diseases (including hypertension with concomitant organ damage), COPD or other chronic respiratory diseases, chronic liver disease, hemoglobinopathies, neurodevelopmental and neurodegenerative disease. Comorbidity burden was assessed using Monoclonal Antibodies Screening Score (MASS^7–22^) that assigned points, as follows: age ≥65 (2 points), BMI ≥35 (1 point), diabetes mellitus (2 points), chronic kidney disease (3 points), cardiovascular disease in a patient ≥55 years (2 points), chronic respiratory disease in a patient ≥55 years (2 points), hypertension in a patient ≥55 years (1 point), and immunocompromised status (3 points).

All consecutive adult patient (age>18) who provided a written informed consent were included in study population. The study was approved by AIFA and National Ethics Committee.

### 2.2 Monoclonal antibody administration

Bamlanivimab/Etesevimab (700 mg/1400 mg) or Casirivimab/Imdevimab (1200 mg/1200mg) were administered by one-hour intravenous infusion and patients were observed for one hour after infusion. Allocation to treatments was pseudo-random, as a criterion of daily alternation (subject to drugs availability) was adopted, and as not many of the measured factors were used to determine which combination to infuse.

Patients who received Bamlanivimab as monotherapy or Sotrovimab were excluded.

### 2.3 Procedures and data collection

Outpatients visits were scheduled at baseline (D1) and at 7 (D7) and 30 (D30) days after infusion. Medical evaluation, vital signs recording, laboratory tests and reports on adverse effects were performed at each visit. If patients missed person visits, they were called by telephone to assess clinical conditions. As a real-life study, due to the high diagnostic demands related to the COVID-19 pandemic, different methods were used to investigate serology and virological parameters according to the laboratory workflow and tests availability.

SARS-COV-2 serology was performed by ELISA detecting anti-SARS-CoV-2 IgG, IgM, and IgA (ENZY-WELL SARS-CoV-2; DIESSE, Diagnostica Senese, Siena, Italy; positive index values ≥1.1), or by two chemiluminescence microparticle assays (CMIA) detecting anti-Nucleoprotein and anti-Spike/RBD IgG (ARCHITECT SARS-CoV-2 IgG, and ARCHITECT SARS-CoV-2 IgG II Quantitative; Abbott Laboratories, Wiesbaden, Germany, respectively). According to the to manufacturer’s instructions, for the two CMIA, Index >1.4 and Binding Antibody Units (BAU)/mL ≥7.1 are considered positive for anti-N and anti-Spike/RBD IgG, respectively.

Semi-quantitative estimation of viral load in NPS was assessed by RT-PCR using DiaSorin Simplexa® COVID-19 Direct platform (DiaSorin, Saluggia, Italy), based on cycle threshold (CT) values of S and ORF1ab genes amplification. Other RT-PCR methods used to verify the presence of SARS-CoV-2 were the Abbott m2000 RealTime System (Abbott Laboratories, Wiesbaden, Germany) and the Cobas® SARS-CoV-2 Test on the fully-automated cobas® 6800 Systems (Roche Diagnostics, Rotkreuz, Switzerland).

Identification of VoCs was conducted by Sanger sequencing of the Spike coding gene on the D1 samples. During the period of Delta variant wave, the RT-PCR Simplexa® SARS-CoV-2 Variants Direct kit (DiaSorin, Saluggia, Italy).was included in the study as rapid method for the qualitative detection and differentiation of the N501Y, E484K, E484Q and L452R mutations.

### 2.4 Outcomes

The primary outcome was defined as time to hospitalization due to development of severe COVID-19 or death from any cause over days 0-30. Secondary outcomes were a) time to hospitalization or death from any cause by day 30, b) time to hospitalization or death from any cause over days 3-30, c) the impact of intervention on CT values change from baseline to D7. The proportion of participants reporting adverse events were also shown as a safety endpoint.

### 2.5 Statistical analysis

Main characteristics of the participants, assessed at D1, were compared by treatment strategy using Mann-Whitney U test for continuous variables, expressed as median (IQR) or χ^2^ test or Fisher’s exact test as appropriate for Categorical variables expressed as numbers and percentages. The effectiveness of the two strategies on the three outcomes was estimated and compared using a weighted pooled logistic regression model which approximates the parameters of a marginal structural Cox model by mean of inverse probability weights. Participants’ follow-up accrued from the date of infusion until the date of hospitalisation, death or date of discharge. Administrative censoring was also applied at 31/12/2021 the date at which the database was frozen. Weights have been calculated using the predicted values from the pooled logistic models for the probabilities of starting BAM/ETE vs. CAS/IMD and those of censoring, respectively. Treatment was fitted as a time-fixed variable and there was no need to account for immortal time bias. Potential informative censoring was controlled for using inverse probability of censoring weights. Unweighted and weighted hazard ratios (HRs) with 95% confidence intervals (CI) were shown. Assumptions regarding the underlying causal link between measured factors are pictured in Figure 1. Unweighted and weighting Kaplan-Meier estimates of the primary outcome stratified by treatment strategy were fitted. Interactions between the intervention and study population strata (type of VoC, vaccination status, level of D1 CT values and period of enrolment) were formally tested by including a multiplicative term in the marginal Cox regression model and adjusted hazard ratios (HRs) with 95% confidence intervals (CI) were shown in a forest plot.

**Figure 1.**
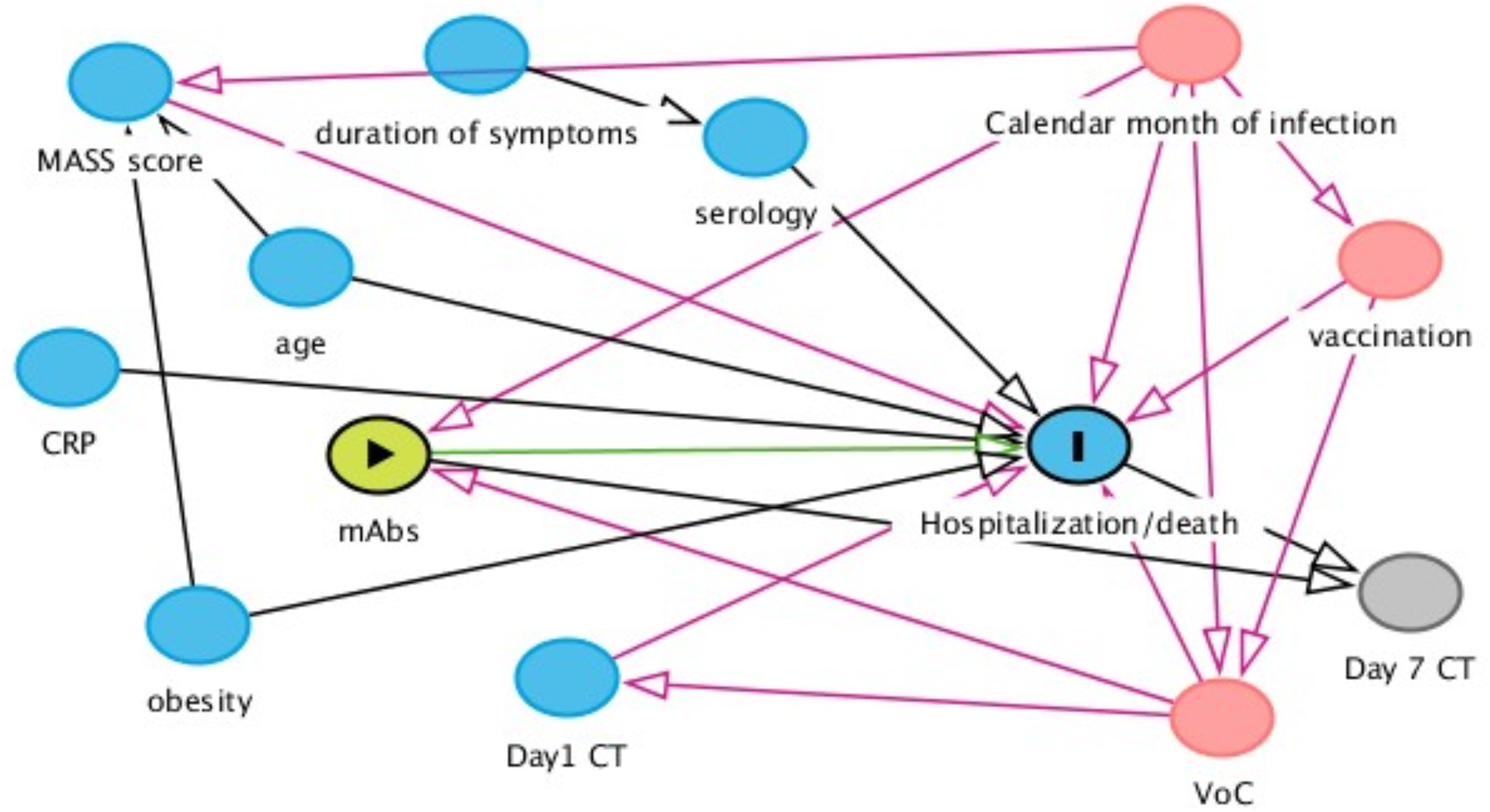
Assumptions regarding the underlying causal link between measured factors. Allocation to BAM/ETE or CAS/IMD was pseudo random as not many of the measured factors were used to determine which combination to use. According to our assumptions, month of infusion and type of VoC were identified as main time-fixed confounders of our comparison of interest. MASS score was an important predictor and used in some of the models to increase efficiency. MASS score assigned points, as follows: age ≥65 (2 points), BMI ≥35 (1 point), diabetes mellitus (2 points), chronic kidney disease (3 points), cardiovascular disease in a patient ≥55 years (2 points), chronic respiratory disease in a patient ≥55 years (2 points), hypertension in a patient ≥55 years (1 point), and immunocompromised status (3 points).

We compared mean CT values at D7 by treatment in analysis of covariance (ANCOVA-model adjusted for D1 CT value, month of enrolment and type of VoC) and we described, using box-plots, median (IQR) of D1 and D7 CT values and its variation.

We also evaluated the associations between D7 CT response (using the cut-offs of 40 for negativity and 25 for reduced viral load) with both the intervention and the primary outcome using a chi-square and Fisher exact test as appropriate.

A descriptive analysis of self-reported side effects was also performed.

A two-sided test of less than 0.05 was considered statistically significant. All statistical analyses were performed using the SAS software, version 9.4 (Carey USA).

## 3. Results

### 3.1 Study Population

From 23^rd^ March 2021 to the 3^rd^ of December 2021, a total of 513 participants were evaluated for receiving mAb treatment and 433 of them (84.4%) were included in the analysis (Figure 2). Briefly, 201 (46%) were female, median age was 63 years (IQR 53-73) and 241 patients (57%) were vaccinated. At baseline, median MASS score was 2 (IQR 0-4) and median time from symptoms onset to D1 was 5 days (IQR 3-6). Alfa (B.1.1.7) variant were identified in 71 participants (22%), Gamma (P.1) in 25 (8%) and Delta (B.1.617.2) in 192 (59%); Beta (B.1.135) and Eta (B.1.525) in one participant each. Negative SARS-COV-2 serology at D1 was detected in 334 (77%) participants; patients receiving CAS/IMD were more likely to have positive serology (34% vs 11%, p<0.001). Among vaccinated patients, mRNA-1273 was used more frequently in patients receiving BAM/ETE, and ChAdOx1 less frequently (p=0.05). Overall, the participants receiving the two treatment strategies appeared to be balanced with respect to key predictors of outcomes as expected under our pseudo-random allocation design. Main characteristics according to the two treatment groups are reported in Table 1.

**Figure 2.**
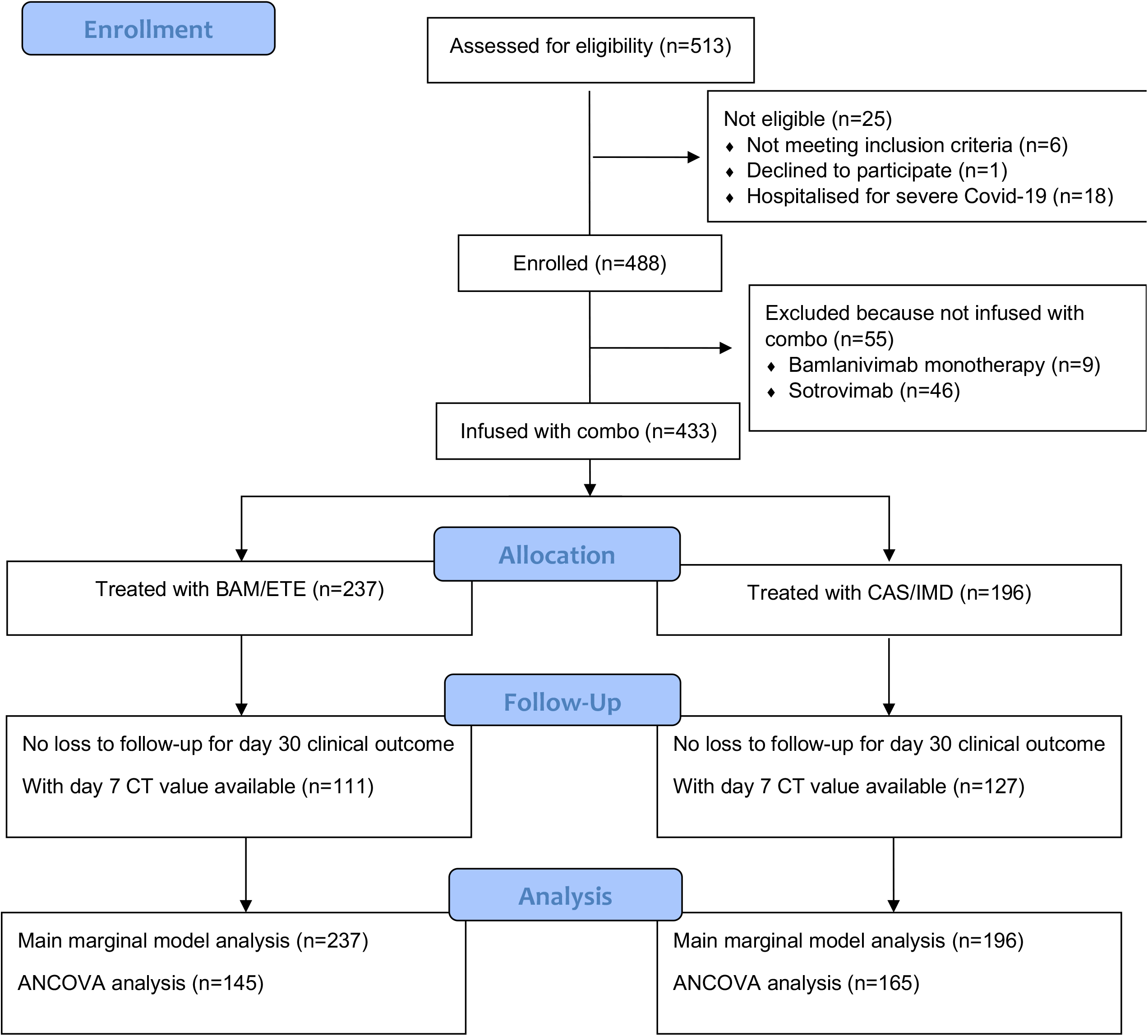
Study flow chart.

**Table 1.** Main characteristics at enrolment by intervention group.

| Characteristics | Regimen started |  |  | Total |
| --- | --- | --- | --- | --- |
|  | BAM/ETE | CAS/IMD | P-value* |  |
|  | N= 237 | N= 196 |  | N= 433 |
| **Gender, n(%)** |  |  | 0.701 |  |
| Female | 112 (47.3%) | 89 (45.4%) |  | 201 (46.4%) |
| **Age, years** |  |  | 0.798 |  |
| Median (IQR) | 63 (53, 73) | 62 (53, 74) |  | 63 (53, 73) |
| **Days from symptoms onset to MAbs infusion** |  |  | 0.624 |  |
| Median (IQR) | 5 (3, 6) | 5 (3, 6) |  | 5 (3, 6) |
| **Comorbidities/risk factors, n(%)** |  |  |  |  |
| Diabetes | 33 (14.0%) | 16 (8.2%) | 0.056 | 49 (11.4%) |
| Severe obesity (BMI>35) | 33 (13.9%) | 20 (10.2%) | 0.240 | 53 (12.2%) |
| Obesity (BMI>30) | 72 (30.4%) | 46 (23.5%) | 0.108 | 118 (27.3%) |
| Hypertension | 97 (41.5%) | 83 (42.6%) | 0.817 | 180 (42.0%) |
| Cardiovascular disease | 42 (17.9%) | 36 (18.5%) | 0.891 | 78 (18.2%) |
| Cerebrovascular disease | 10 (4.3%) | 10 (5.1%) | 0.669 | 20 (4.7%) |
| Chronic respiratory disease | 37 (15.7%) | 34 (17.3%) | 0.642 | 71 (16.4%) |
| Renal impairment | 4 (1.7%) | 1 (0.5%) | 0.250 | 5 (1.2%) |
| Neurologic disease | 7 (3.0%) | 9 (4.6%) | 0.378 | 16 (3.7%) |
| Autoimmune disease | 24 (10.2%) | 21 (10.7%) | 0.865 | 45 (10.4%) |
| Neoplasms | 19 (8.1%) | 15 (7.7%) | 0.879 | 34 (7.9%) |
| Hematologic disease | 13 (5.5%) | 13 (6.7%) | 0.623 | 26 (6.0%) |
| Immunodeficiency | 11 (4.9%) | 9 (4.7%) | 0.924 | 20 (4.8%) |
| **Vital signs at baseline** |  |  |  |  |
| SpO<sub>2</sub>, median (IQR) | 97 (96, 98) | 97 (96, 98) | 0.289 | 97 (96, 98) |
| Fever (>37.5°C), n(%) | 12 (5.1%) | 14 (7.3%) | 0.342 | 26 (6.1%) |
| BMI, median (IQR) | 26.67 (23.71, 31.89) | 25.92 (23.10, 30.12) | 0.079 | 26.23 (23.46, 31.22) |
| **Laboratory values, median (IQR)** |  |  |  |  |
| Ferritin, ng/ml | 161.5 (68.00, 274.0) | 179.0 (110.0, 313.0) |  | 173.0 (81.00, 296.0) |
| C-reactive protein, mg/dl | 1.33 (0.52, 3.19) | 1.20 (0.49, 2.42) |  | 1.27 (0.50, 2.83) |
| Lymphocytes, /uL | 1210 (850.0, 1600) | 1160 (880.0, 1530) |  | 1180 (870.0, 1560) |
| **Baseline SARS-COV-2** |  |  |  |  |
| **Serology, n(%)** |  |  | <.001 |  |
| Positive | 26 (11.0%) | 66 (33.7%) |  | 92 (21.2%) |
| Negative | 207 (87.3%) | 127 (64.8%) |  | 334 (77.1%) |
| Unknown | 4 (1.7%) | 3 (1.5%) |  | 7 (1.6%) |
| **Vaccination, n(%)** |  |  | 0.077 |  |
| Yes (partly or fully) | 140 (60.6%) | 101 (52.1%) |  | 241 (56.7%) |
| **Vaccine type, n(%)** |  |  | 0.050 |  |
| BNT162b2 | 75 (68.8%) | 65 (68.4%) |  | 140 (68.6%) |
| mRNA-1273 | 18 (16.5%) | 6 (6.3%) |  | 24 (11.8%) |
| ChAdOx1 | 8 (7.3%) | 20 (21.1%) |  | 28 (13.7%) |
| Ad26.COV2.S | 8 (7.3%) | 4 (4.2%) |  | 12 (5.9%) |
| Other/unknown | 31 (22.1%) | 6 (5.9%) |  | 37 (15.4%) |
| **SARS-COV-2 variant, n(%)** |  |  | 0.886 |  |
| B.1.1.7/Alpha | 34 (22.5%) | 37 (21.4%) |  | 71 (21.9%) |
| P.1/Gamma | 14 (9.3%) | 11 (6.4%) |  | 25 (7.7%) |
| B.1.617.2/Delta | 87 (57.6%) | 105 (60.7%) |  | 192 (59.3%) |
| Other VoC | 1 (0.7%) | 1 (0.6%) |  | 2 (0.6%) |
| Not done | 15 (9.9%) | 19 (11.0%) |  | 34 (10.5%) |
| **Baseline CT, mean (SD)** | 21.01 ± 6.46 | 20.14 ± 6.09 |  | 20.56 ± 6.28 |
| **MASS score, median (IQR)** | 2 (0, 4) | 2 (0, 3) | 0.104 | 2 (0, 4) |
| **Enrolled after June 2021, n(%)** | 172 (72.6%) | 127 (64.8%) | 0.082 | 299 (69.1%) |

\*Chi-square or Mann-Whitney test as appropriate
Notes: BAM/ETE, bamlanivimab/etesevimab; CAS/IMD, casirivimab/imdevimab; IQR, interquartile range; BMI, body mass index; SpO<sub>2</sub>, peripheral oxygen saturation; MASS, Monoclonal Antibodies Screening Score

### 3.2 Primary endpoint

COVID19-related hospitalization or death from any cause occurred in 19 participants: 15 patients in the BAM/ETE group (6.3%) and 4 patients in CAS/IMD group (2%) (Table 2). Two deaths were observed, both in patients treated with BAM/ETE experiencing COVI-19 clinical progression. The majority of the events occurred before D7. In the weighted Kaplan-Meier analysis there was greater evidence for a difference in risk by intervention group (>10% for BAM/ETE) than in the unweighted (Figure 3A/3B).

**Table 2.** Main outcomes by intervention groups.

| Outcomes | Regimen started |  |  |  |
| --- | --- | --- | --- | --- |
|  | BAM/ETE | CAS/IMD | P-value* | Total |
|  | N= 237 | N= 196 |  | N= 433 |
| *Primary Outcome<sup>a</sup>, n(%)* | 15 (6.3%) | 4 (2.0%) | 0.030 | 19 (4.4%) |
| *<sup>1st</sup> Secondary Outcome<sup>b</sup>, n(%)* | 21 (8.9%) | 8 (4.1%) | 0.048 | 29 (6.7%) |
| *<sup>2nd</sup> Secondary Outcome<sup>c</sup>, n(%)* | 9 (3.8%) | 2 (1.0%) | 0.068 | 11 (2.5%) |

Notes: BAM/ETE, bamlanivimab/etesevimab; CAS/IMD, casirivimab/imdevimab
<sup>a</sup>Patients with COVID-19 related hospitalization or death for any cause over days 0-30
<sup>b</sup>Patients with hospitalization or death for any cause over days 0-30
<sup>c</sup>Patients with hospitalization or death for any cause over days 3-30
\* Chi-square

**Figure 3.**
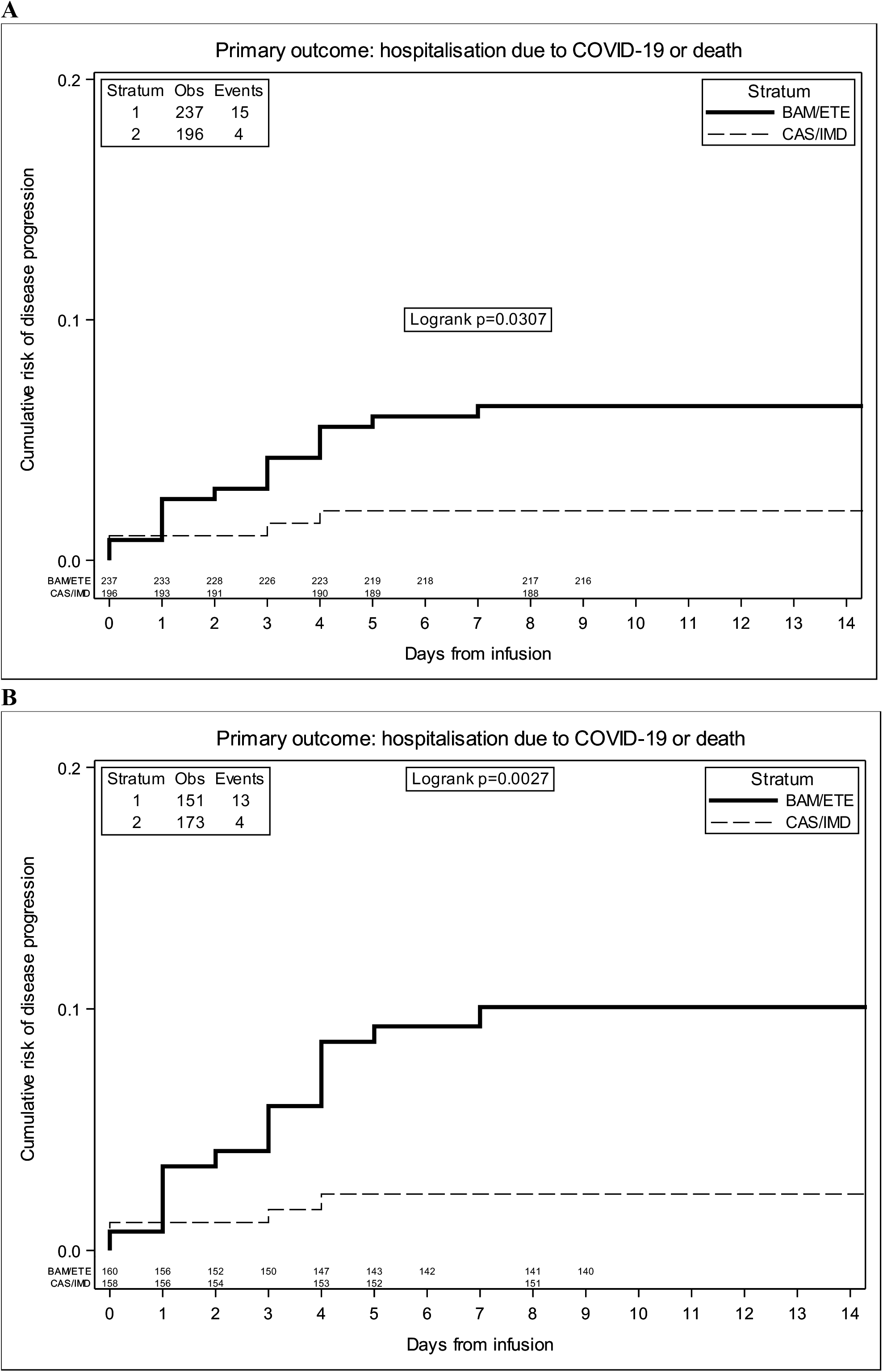
Unweighted (A) and Weighted (B) Kaplan Meier curves of time to primary endpoint by intervention group-weights include month of enrolment and type of VoC.

The relative hazard of COVID-19-related hospitalization or death for CAS/IMD compared to BAM/ETE was 0.28 (95%CI 0.09-0.85; p=0.024, Table 3A).

**Table 3.** Relative Hazard from fitting a marginal model using BAM/ETE as the comparator group for primary (A) and secondary endpoints (B-C).

**A. RH from fitting a marginal model for primary endpoint**
|  | Unadjusted and adjusted marginal relative hazards of hospitalization/death |  |  |  |
| --- | --- | --- | --- | --- |
|  | Unadjusted HR (95% CI) | p-value | Adjusted** HR (95% CI) | p-value |
| BAM/ETE | 1.00 |  | 1.00 |  |
| CAS/IMD | 0.32 (0.10, 0.95) | 0.041 | 0.28 (0.09, 0.85) | 0.024 |

**B. RH from fitting a marginal model for 1st secondary endpoint**
|  | Unadjusted and adjusted marginal relative hazards of hospitalization/death |  |  |  |
| --- | --- | --- | --- | --- |
|  | Unadjusted HR (95% CI) | p-value | Adjusted** HR (95% CI) | p-value |
| BAM/ETE | 1.00 |  | 1.00 |  |
| CAS/IMD | 0.45 (0.20, 1.02) | 0.056 | 0.40 (0.17, 0.91) | 0.029 |

**C. RH from fitting a marginal model for 2nd secondary endpoint**
|  | Unadjusted and adjusted marginal relative hazards of hospitalization/death |  |  |  |
| --- | --- | --- | --- | --- |
|  | Unadjusted HR (95% CI) | p-value | Adjusted** HR (95% CI) | p-value |
| BAM/ETE | 1.00 |  | 1.00 |  |
| CAS/IMD | 0.26 (0.06, 1.22) | 0.088 | 0.22 (0.05, 1.00) | 0.051 |

Notes: BAM/ETE, bamlanivimab/etesevimab; CAS/IMD, casirivimab/imdevimab
<sup>a</sup>Patients with COVID-19 related hospitalization or death for any cause over days 0-30
<sup>b</sup>Patients with hospitalization or death for any cause over days 0-30
<sup>c</sup>Patients with hospitalization or death for any cause over days 3-30
\*\* adjusted for calendar month, MASS score and VoCs

### 3.3 Secondary endpoints

Hospitalization or death from any cause by day 30 occurred in 29 participants (9% in BAM/ETE vs. 4% in CAS/IMD). Overall, 18 of these secondary events (62%) occurred within day three after infusion (Table 2).

Table 3B and 3C, showed the weighted relative hazard ratio for secondary endpoints.

### 3.4 Effect measure modification analysis

The analysis by subsets carried no evidence that the effect of the intervention was different across a number of stratification variables for the primary endpoint (p-values for interaction >0.18, Figure 4). The effect of the intervention appeared attenuated in participants with the Delta VoC but with large uncertainty around these subgroup estimates.

**Figure 4.**
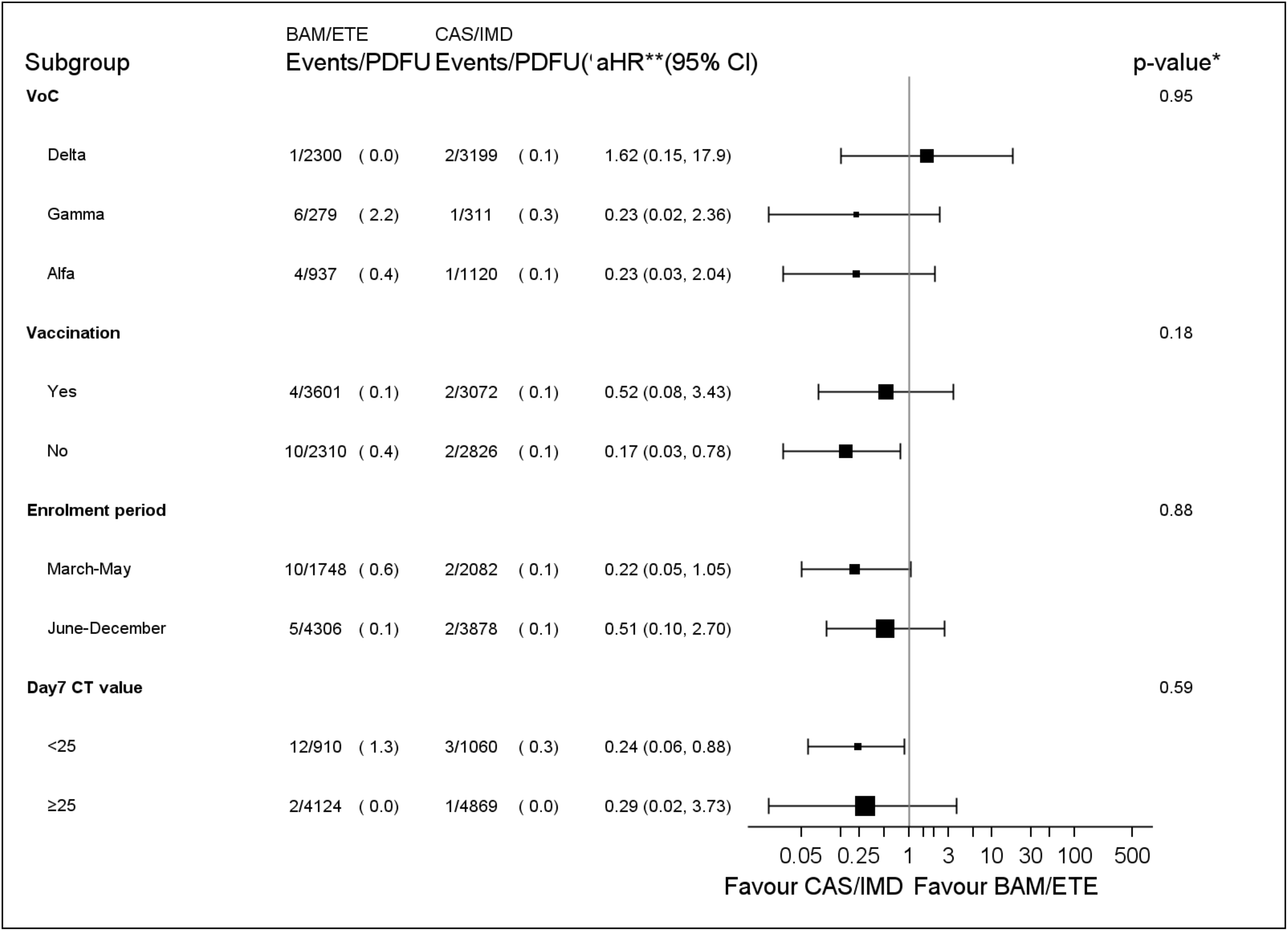
Forest plot of the effect of the intervention by subsets - Primary endpoint. *p-value corresponds to the test for interaction between intervention and each subgroup unadjusted for multiplicity **aHR1– (vaccination and CT strata) adjusted for calendar month and VoC; **aHR2 – (VoC stratum) adjusted for MASS score because of a positivity issue leading to abnormally high weights in the stratum of participants infected with the Delta VoC; **aHR3 – (enrolment period stratum) adjusted for MASS score and VoC PDFU= person days of follow-up

### 3.5 Analysis of covariance

Median (IQR) of CT at D1 and D7 and CT increase between D1 and D7 are shown in Figure 5A/5B. We found no evidence for a difference in D7 CT mean values comparing CAS/IMD vs. BAM/ETE (+0.17, 95%: −1.41; +1.74, p=0.83). Of note, proportions of participants with a negative SARS-COV-2 RT-PCR in NPS and with a high viral load at D7 were 26.1% in BAM/ETE vs. 25.8% in CAS/IMD (p=0.94) and 22.1% in BAM/ETE vs. 18.9% in CAS/IMD (p=0.46), respectively. Among patients who experienced primary outcome, none showed a negative SARS-COV-2 RT-PCR in NPS (p=0.009) and 82.4% showed still high viral load (p<0.001) on D7.

**Figure 5.**
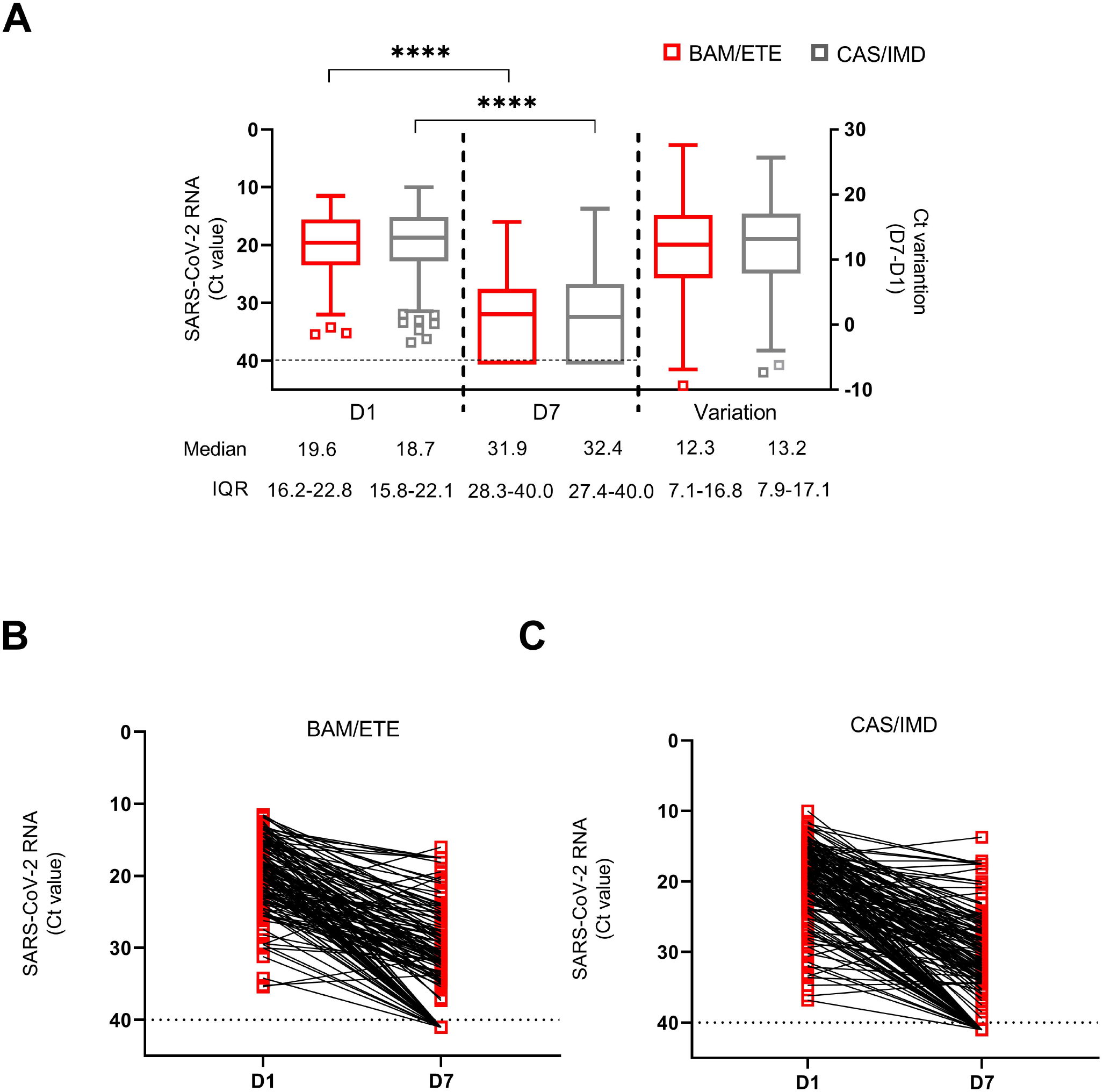
SARS-CoV-2 RNA levels at D1 and D7 in patients treated with CAS/IMD and BAM/ETE. **(A)** Box-and-whiskers plot showing the comparison of viral loads detected at D1 and D7 in patients treated with BAM/ETE (n=170 and 154, respectively) or CAS/IMD (n=183 and 174, respectively), and the variation of RNA levels observed between the two time-points by intervention (BAM/ETE, n=145; CAS/IMD, n=165). Viral RNA levels are expressed as CT of Orf1ab gene amplification. Median CT values and IQR are shown. Statistical analysis of the comparisons between treatment groups was performed by Mann–Whitney test: D1, p=0.29; D7, p=0.87; and Variation, p=0.71. CT values at D1 and D7 within each group were compared using paired Wilcoxon sign-rank test, ****p<0.0001. **(B-C)** Spaghetti-plot showing the Orf1ab CT values measured at D1 and D7 in each patient treated with BAM/ETE (n= 145, B) or CAS/IMD (n=165, C). Horizontal dashed line represents the limit of detection (CT: 40.0), values ≥40 are considered negative.

### 3.6 Adverse events

Thirty-one participants reported adverse events; a breakdown of the most common adverse events reported is described in Table 4. Overall, 19 events (10%) in the CAS/IMD group and 6 events (3%) in the BAM/ETE group were considered related to mAb infusion (p=0.001). One patient only (0.2%) reported severe dyspnea requiring hospitalization: this event was not considered by the investigators to be related to treatment.

**Table 4.** Proportions of participants reporting specific adverse events by intervention group.

| Characteristics | Regimen started |  |  | Total |
| --- | --- | --- | --- | --- |
|  | BAM/ETE | CAS/IMD | P-value* |  |
|  | N= 237 | N= 196 |  | N= 433 |
| ***Adverse events, n(%)*** |  |  | 0.064 |  |
| Mild | 8 (3.4%) | 15 (7.7%) |  | 23 (5.3%) |
| Moderate | 0 (0.0%) | 7 (3.6%) |  | 7 (1.6%) |
| Severe | 1 (0.4%) | 0 (0.0%) |  | 1 (0.2%) |
| Drug related | 6 (2.5%) | 19 (9.7%) | 0.001 | 25 (5.8%) |
| ***Individual events, n(%)*** |  |  |  |  |
| Arthomyalgia | 1 (0.4%) | 2 (1.0%) | 0.455 | 3 (0.7%) |
| Asthenia | 0 (0.0%) | 0 (0.0%) |  | 0 (0.0%) |
| Chills | 0 (0.0%) | 3 (1.5%) | 0.056 | 3 (0.7%) |
| Diarrhea | 1 (0.4%) | 1 (0.5%) | 0.893 | 2 (0.5%) |
| Fever | 4 (1.7%) | 17 (8.7%) | <.001 | 21 (4.8%) |
| Nausea | 0 (0.0%) | 4 (2.0%) | 0.027 | 4 (0.9%) |
| Sight decrease | 0 (0.0%) | 1 (0.5%) | 0.271 | 1 (0.2%) |
| Headache | 0 (0.0%) | 1 (0.5%) | 0.271 | 1 (0.2%) |
| Dyspnea | 1 (11.1%) | 1 (4.5%) | 0.506 | 2 (6.5%) |
| Abdominal pain | 1 (11.1%) | 0 (0.0%) | 0.118 | 1 (3.2%) |
| Hematoma | 0 (0.0%) | 0 (0.0%) |  | 0 (0.0%) |
| Paraesthesia | 1 (0.4%) | 0 (0.0%) | 0.363 | 1 (0.2%) |
| Itch | 0 (0.0%) | 0 (0.0%) |  | 0 (0.0%) |
| Skin rash | 0 (0.0%) | 2 (1.0%) | 0.119 | 2 (0.5%) |
| Dizziness | 0 (0.0%) | 0 (0.0%) |  | 0 (0.0%) |
| Vomit | 0 (0.0%) | 1 (0.5%) | 0.271 | 1 (0.2%) |

\*Chi-square or Mann-Whitney test as appropriate
Notes BAM/ETE, bamlanivimab/etesevimab; CAS/IMD, casirivimab/imdevimab; IQR, interquartile range; BMI, body mass index; SpO2, peripheral oxygen saturation

## 4. Discussion

To our knowledge, this analysis is the first real-life comparison of two routinely used anti-SARS-CoV-2 mAbs by a trial emulation methodology using observational data collected from outpatients with early mild-to-moderate COVID-19 and at high-risk for disease progression. In an evolving scenario of SARS-CoV-2 variants from Alpha to Delta, we found that patients receiving Casirivimab/Imdevimab had a 72% lower risk of COVID-19 related-hospitalization or death from any cause than patients receiving Bamlanivimab/Etesevimab. The greater benefit of Casirivimab/Imdevimab was evident also excluding patients who experienced failure by day2 (a time window in which events cannot be ascribed to lack of treatment effect), with a confirmed 78% risk reduction.

Overall, we found no evidence that the magnitude of the difference of the effect between mAb interventions on the risk of clinical outcomes was different in specific subsets of the study population. Nevertheless, the effect of Casirivimab/Imdevimab appeared to be larger in subgroups with the highest known risk of disease progression: patients with higher baseline viral load^23^, unvaccinated^24–25^ and those enrolled before June 2021, when the target population included people with higher risk of severe outcome^26^.

On the other hand, there was larger uncertainty around the hazard ratio comparing interventions in the subgroup of patients infected with the Delta VoC with a 95% CI not excluding superiority of Bamlanivimab/Etesevimab vs. Casirivimab/Imdevimab. This finding is consistent with *in vitro* observed retained activity of Bamlanivimab/Etesevimab^15–27^ against Delta variant (B.1617.2, non-AY.1/AY.2), and supports the recommended use of both Bamlanivimab/Etesevimab and Casirivimab/Imdevimab in settings of elevated Delta VoC prevalence.

To our knowledge, this is the first analysis from real-life data evaluating virological response to mAb treatment. It has been suggested that mAbs may act as antiviral neutralizing agents through multiple mechanisms, such as targeting free virus and virally infected cells^28^. Significant decrease in viral load was described in RCT^2–4^ for the two mAbs combinations analyzed in this study, but relation between virological and clinical outcomes remains uncertain^29^. Interestingly, our analysis showed no difference in terms of viral load reduction from D1 to D7 between Casirivimab/Imdevimab and Bamlanivimab/Etesevimab, but also displayed a very strong association between clinical and virological outcomes, suggesting that patients developing severe disease also failed in viral clearing. However, viral load was measured after the occurrence of all clinical events, so it is difficult to determine how much clinical outcome was mediated by the virological response or whether lack of virological clearance was actually a consequence of the clinical picture.

Finally, our data confirmed safety and tolerability of these two mAb combinations in a real-life unselected population.

Our analysis has some limitations. First, due to the observational nature of the study conducted in a single COVID health care center and to the lack of a randomized design, confounding bias cannot be ruled out. Further, eligibility criteria changed over time concurrently with the advent of Delta wave and with a wider use of Bamlanivimab/Etesevimab, due to available supplies. However, results were similar after controlling for MASS score in the regression models. Moreover, the lack of an early measure of CT (e.g. at D3) prevented us from investigating viral load as a potential mediator. Finally, the study was conducted before the emergence of Omicron B.1.1.529 VoC, which is going to subvert previous assessment about mAbs treatment as several *in vitro* studies suggest that both Bamlanivimab/Etesevimab and Casirivimab/Imdevimab did not retain a remarkable activity against Omicron^16–30–31–32^. Despite this, even today a proportion of illnesses and consequent hospitalizations are still due to the VoCs different from Omicron, and so knowledge of comparative data between available mAbs is still crucial for optimizing treatment in pandemic times.

## Data Availability

All data produced in the present work are contained in the manuscript

## Authors’Contributions

AA, VM conceptualized and designed the study. VM and IM wrote the protocol. VM, FC and ACL wrote the first draft of the manuscript and referred to appropriate literature. ACL was also the main responsible person for formal data analysis. AA, ACL, VM, AC, FC, CC and SL conceived, supervised the study and contributed to data interpretation. CC and PP were responsible for data curation. AV, IM, ARG, SC, EG, EN, FV revised the manuscript content, reviewed and edited the manuscript. EL, FC, CC and ARG performed all virological test. IM, AV, SL, SR enrolled participants. All authors agreed with and approved the final version of the manuscript.

## Acknowledgements

The authors gratefully acknowledge nurse staff, all the patients and all members of the Outpatient clinic for MAbs administration and of laboratory:

Chiara Agrati, Andrea Antinori, Tommaso Ascoli-Bartoli, Francesco Baldini, Bartolini Barbara, Rita Bellagamba, Giulia Berno, Nazario Bevilacqua, Giulia Bonfiglio, Licia Bordi, Marta Camici, Rita Casetti, Concetta Castilletti, Stefania Cicalini, Claudia Cimaglia, Francesca Colavita, Angela Corpolongo, Alessandro Cozzi Lepri, Federico De Zottis, Angela D’Urso, Lavinia Fabeni, Massimo Francalancia, Marisa Fusto, Roberta Gagliardini, Letizia Giancola, Giuseppina Giannico, Emanuela Giombini, Enrico Girardi, Giulia Gramigna, Elisabetta Grilli, Susanna Grisetti, Cesare Ernesto Maria Gruber, Giuseppina Iannicelli, Eleonora Lalle, Simone Lanini, Gaetano Maffongelli, Andrea Mariano, Ilaria Mastrorosa, Giulia Matusali, Valentina Mazzotta, Silvia Meschi, Annalisa Mondi, Emanuele Nicastri, Sandrine Ottou, Claudia Palazzolo, Jessica Paulicelli, Carmela Pinnetti, Pierluca Piselli, Maria Maddalena Plazzi, Alessandra Oliva, Silvia Rosati, Martina Rueca, Alessandra Sacchi, Giuseppe Sberna, Laura Scorzolini, Francesco Vaia, Alessandra Vergori, Serena Vita.

## Access to data

Anonymized participant data will be made available upon reasonable requests directed to the corresponding author. Proposals will be reviewed and approved by investigator, and collaborators on the basis of scientific merit. After approval of a proposal, data can be shared through a secure online platform after signing a data access agreement.

